# “We have been so patient because we know where we are coming from” Exploring the acceptability and feasibility of a mobile electronic medical record system designed for community-based antiretroviral therapy in Lilongwe, Malawi

**DOI:** 10.1101/2024.04.23.24306213

**Authors:** Christine Kiruthu-Kamamia, Astrid Berner-Rodoreda, Gillian O’Bryan, Odala Sande, Jacqueline Huwa, Agnes Thawani, Hannock Tweya, Wim Groot, Milena Pavlova, Caryl Feldacker

## Abstract

**Background:** Mobile health (mHealth) is reshaping healthcare delivery, especially in HIV management. The World Health Organization advocates for mHealth to provide healthcare workers (HCWs) with real-time data, enhancing patient care. However, in Malawi’s Lighthouse Trust antiretroviral therapy (ART) clinic, the nurse-led community-based ART (NCAP) program faces hurdles with data management due to lack of access to electronic medical records systems (EMRS) in the community setting. EMRS is not typically available in differentiated service delivery settings where reliable power and internet are often unavailable. We used human-centered design (HCD) processes to create a mobile EMRS prototype, the Community-based ART Retention and Suppression (CARES) app. We explore progress to simplify workflow for HCWs and improve client care.

**Methods:** To evaluate the CARES app’s feasibility and acceptability among NCAP HCWs, we conducted in-depth interviews among 15 NCAP HCWs. We used a rapid qualitative analysis approach guided by the extended Technology Acceptance Model. The study complied with the Consolidated Criteria for Reporting Qualitative Research (COREQ).

**Results:** As a likely result of HCD, HCWs demonstrated high expectations for the CARES app to improve healthcare delivery and data management. However, challenges such as app performance, data integration, and system navigation were significant barriers to acceptance or feasibility. Despite challenges, HCWs remained optimistic about the potential for CARES to enhance NCAP clinical decision-making and data flow. HCWs emphasized the need for continuous training and stakeholder engagement, improved infrastructure, data security protections, and establishing the CARES app and EMRS integration to facilitate CARES’ long-term success at scale.

**Conclusion:** The study’s findings underscore the importance of HCD for mHealth buy-in. As HCWs were invested in CARES success, they remained optimistic that the app could enhance NCAP services if user experience and app performance improved. Incorporation of HCW feedback would help deliver beyond the promise of CARES.

## Introduction

Mobile health (mHealth), the use of mobile devices to improve healthcare delivery [1], has become a pivotal force of change in healthcare, including for HIV management [2–5]. Recognizing the potential of technological advancements, the World Health Organization (WHO) emphasizes using innovative mHealth technologies to optimize person-centered service delivery, ensuring HCWs have the necessary data at the point of care [6]. In low-resource settings, mHealth innovations have been shown to improve HIV treatment outcomes, including enhanced antiretroviral treatment (ART) adherence, retention, and viral load suppression [7–9].

Notably, HIV remains a pervasive global issue, particularly in sub-Saharan Africa, with countries like Malawi experiencing some of the world’s highest HIV prevalence rates. The WHO advocates the adoption of differentiated service delivery (DSD) models in HIV care, which provide tailored care to people living with HIV regardless of their age, socioeconomic status, background, viral load levels, and stability on ART [10]. Community-based DSD models shift stable ART clients from crowded clinics to community care [10,11], like the nurse-led community ART program (NCAP) piloted at the Lighthouse Trust ART clinic in Malawi [12]. Lighthouse Trust is the largest ART provider in Malawi, providing care to over 65,000 clients alive on ART [13,14]. NCAP is a community DSD that has been operational for nearly eight years at Lighthouse Trust [12] . Community ART distribution enables patients to refill ART closer to their homes through mobile clinics or community meeting points, reducing travel times for the client and decongesting healthcare facilities [11].

Providing quality care requires a robust, comprehensive monitoring and evaluation (M&E) system [15,16]. This is often more complex, but not less necessary, in a community ART setting. In community DSD, HCWs typically rely on paper-based reporting, which can increase the risk of loss-to-follow-up and impede viral load (VL) monitoring[17–19]. After NCAP was piloted, workers and clients responded favorably, and clients preferred receiving ART in the community DSD model rather than in a facility. However, workers found NCAP M&E challenges due to the lack of an electronic medical record system (EMRS), which eases the workload and streamlines reporting [12]. In Malawi, the EMRS was developed to overcome the lack of reliable data and inadequate use of data in healthcare planning [20] . To date, EMRS is only available in static hospitals or health facilities with stable connectivity. Community settings lack such infrastructure [20] . This poses a critical challenge to EMRS expansion in Malawi. mHealth can be used to fill this gap by connecting with the EMRS system to strengthen ART patient care and management. Interoperating mHealth with EMRS holds promise to expand EMRS reach, thereby streamlining data capture, management, and analytics to enhance efficiency [21–23]. Interoperability may also help ensure continuous patient monitoring and support, regardless of location, leading to more informed decision-making and improved care outcomes.

Lighthouse Trust, the International Training and Education Centre for Health, and Medic collaborated to develop a tablet-based EMRS app for NCAP: Community-based ART REtention and Suppression (CARES). The CARES app prototype is designed explicitly for the NCAP at Lighthouse Trust and the Malawi EMRS. It emulates the functionality and features of the EMRS with built-in prompts and alerts, ensuring compliance with national ART guidelines and enhancing the quality of patient care. We used participatory human-centered design (HCD) processes to design the CARES app, working iteratively with NCAP workers to launch the CARES prototype [24]. For Lighthouse clients, CARES operates like EMRS, mimicking the flow and content of routine visits for clients in the clinic setting. The CARES app and the HCD process utilized for its development were detailed previously.

The aim of this study was to qualitatively explore the acceptability and feasibility of the CARES prototype among Lighthouse Trust ART workers who routinely interact with the CARES app or its data. This study took place during the CARES prototype testing and optimization phase. In-depth interviews (IDIs) with key healthcare workers (HCWs) involved in CARES aimed to identify facilitators and barriers to the CARES prototype from the perspective of healthcare services delivery. Data collection and analysis were informed by the Technology Acceptance Model (TAM) information systems theory [25], which aims to explain user behavior regarding technology acceptance, focusing on perceived usefulness and perceived ease of use as fundamental determinants of technology adoption. Understanding the perspectives of HCW users on the CARES prototype app is essential for understanding the intervention’s challenges, opportunities, and potential impact in enhancing ART care delivery and, ultimately, client outcomes. Our findings can also be used to critically assess the applicability of TAM to developing an app for improved community services. Overall, we hope our research will inform both CARES improvements and the development and implementation of other future mobile EMRS interventions for community ART programs in Malawi and beyond and critically assess the applicability of TAM to developing an app for improved community services.

## Methodology

### Design

A qualitative design, using in-depth interviews to ascertain HCW perspectives and experience in using the CARES app. This study’s reporting follows the Consolidated Criteria for Reporting Qualitative Studies (COREQ) guidelines[26]. (see S1 Appendix).

### Setting

Lighthouse Trust operates five centers of excellence clinics across the country. In urban Lilongwe, Lighthouse Trust operates two flagship clinics in urban Lilongwe, Malawi, with a combined >37,000 ART patients[13,14,27]. In addition, Lighthouse Trust also supports care at seven peri-urban satellite sites in Lilongwe District. Lighthouse Trust uses the Malawi Ministry of Health (MoH) real-time, point-of-care EMRS to guarantee compliance with national ART guidelines, enhance patient care and program outcomes, and simplify reporting across all facilities. As with other MoH clinics, the EMRS at each Lighthouse facility operates independently and does not share patient data across different clinics. The NCAP services are offered to clients from Lighthouse Trust flagship clinics, Lighthouse Trust (LT) and Martin Preuss Centre (MPC), and seven satellite health facilities in Lilongwe. To date, the CARES prototype has been utilized only for clients at LT and MPC clinics.

Since 2016, Lighthouse Trust’s NCAP community DSD has provided ART in the community to stable ART clients over 18 who have been on first or second-line treatment for at least six months and are virally suppressed within Lighthouse’s catchment area [12,24]. NCAP offers monthly support group services, supplying 3–6-month ART refills and facilitating specialist referrals when necessary. Previously managed with an ODK-based tool, data for NCAP visits are manually transferred to EMRS, a process CARES aims to streamline. CARES prototype addresses several ODK limitations, including decision support, integrated care, lab management, and reducing clerical workload.

### Study Population

We conducted in-depth interviews (IDIs) of HCWs who used the CARES prototype while providing care in the community and those who worked on the technical side, managing, and processing the CARES app’s data. All HCWs involved with NCAP were approached to participate in the interviews, and all agreed to participate in the study. The HCWs interviewed included NCAP nurses, their managers, and all M&E staff involved in NCAP data management, IT, and research. All the nurses provide direct services to NCAP clients in the community. The community health services (CHS) manager and deputy manager oversee the NCAP program and supervise the nurses. The data officer is responsible for manually entering NCAP data into the EMRS and downloading a list of NCAP clients who missed their appointments. The IT manager is responsible for hosting the electronic data collection tools for NCAP, designing them, and managing the data as needed. The data manager supports data extraction and reporting for the CHS team and develops NCAP data collection tools. The research intern and research manager support the implementation of the CARES project. By interviewing all HCWs involved in the NCAP program, we anticipated reaching a natural point of data saturation.

### Developing CARES using a human-centered design (HCD) approach

Medic, I-TECH, and Lighthouse Trust collaborated as a multidisciplinary team to co-create the CARES prototype app. CARES app was created utilizing the Community Health Toolkit (CHT)[28], an open-source project developed by Medic. HCD was used to engage HCWs consistently, incorporate iterative feedback, and create contextually relevant technology. CARES functionality is described in detail elsewhere[24]. In brief, CARES is a patient management application for ART that prioritizes offline functionality and is designed for use in areas with limited internet connectivity – like NCAP. CARES aims to extend the reach of the Malawi MoH EMRS. In the static facility setting, CARES was designed to assist HCWs in gathering ARV drugs for all clients they expect to encounter in the community. In the NCAP setting, CARES aimed to benefit HCWs by offering them a mobile EMRS-like app, facilitating comprehensive client reviews, and aiding decision-making with alerts for various concerns. It also allows for the enrollment of new clients in the NCAP program. After each visit, CARES aimed to automatically sync data to point-of-care EMRS, eliminating the need for manual data entry and ensuring efficient record-keeping.

The CARES prototype was designed using the HCD approach, with Medic developers having routine discussions with NCAP providers over 12 months to increase the CARES app’s usability and acceptability in the NCAP setting. This lengthy but highly participatory approach prioritized understanding of diverse end-user needs, minimizing wasted resources on irrelevant features. Users included clinicians, nurses, M&E teams, data managers, and NCAP leadership. The feedback sessions, occurring weekly or twice a week, with both theory and hands-on activities, involved developers engaging providers and collaborating closely with a diverse team to tailor CARES system requirements to the local context, with iterative improvements guided by analyzing user feedback and system usage patterns[29].

### Theoretical Model

Analysis of IDIs was guided by the extended Technology Acceptance Model (TAM2)[30]. We present the results according to the TAM2 framework, including the following constructs: Perceived Usefulness, Perceived Ease of Use, and external factors that influence the behavioral intention to use new technology. TAM2 is based on the original TAM, which identifies two distinct attitudes that serve as predictors for adopting a new information system: perceived ease of use and usefulness[30] . TAM2 (Fig. 1) expands the TAM theory by incorporating additional constructs, social influence processes, and cognitive instrumental processes that influence user acceptance through perceived usefulness. Social influence processes explain the relationships among social factors, including subjective norm, voluntariness, and image, which affect a person’s views on the usefulness of technology and their willingness to use it. The cognitive instrumental processes refer to individuals evaluating how well a system or technology aligns with their job relevance, result demonstrability, output quality, and perceived ease of use, all of which contribute to their perceived usefulness. The constructs are defined in Table 1. In other words, the cognitive instrumental process is a practical thinking process where an individual considers how a technology or system fits into their work.

**Fig. 1:**
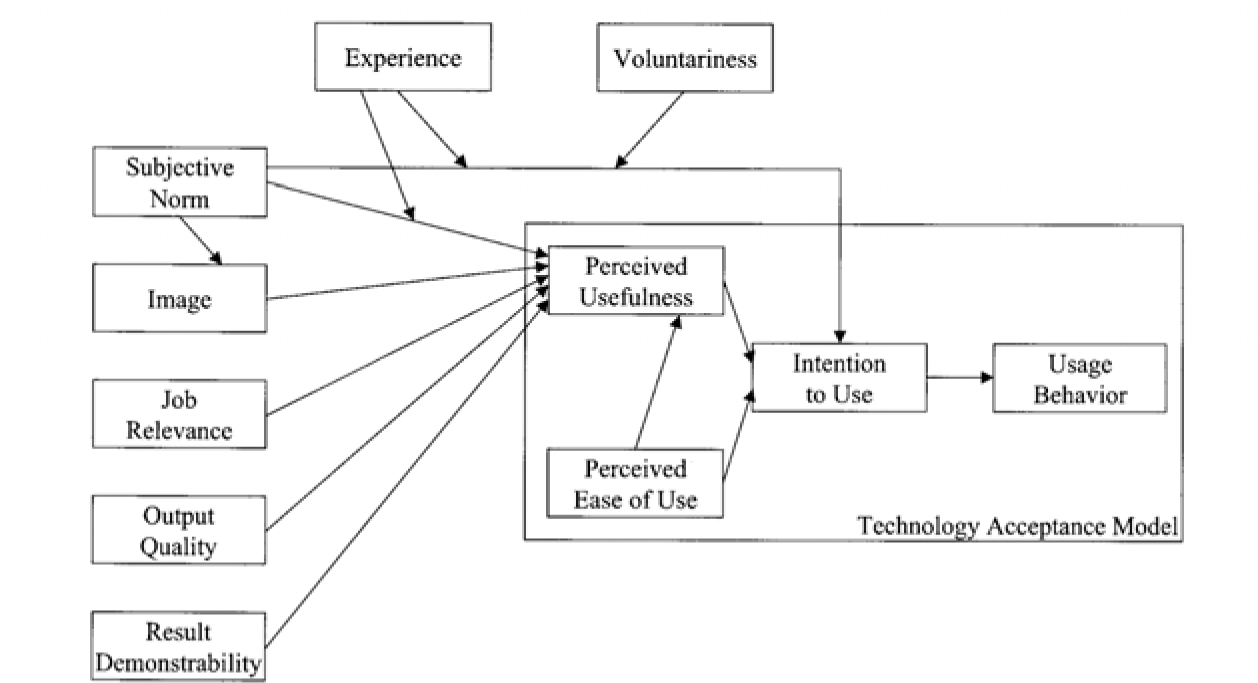
TAM2 proposed by Venkatesh and Davis[30]

**Table 1:** Definitions

| Construct | Definition |
| --- | --- |
| Feasibility | The feasibility of relevant stakeholders to implement the intervention[31]. |
| Acceptability | The acceptability of the intervention to clients and HCWs[31]. |
| Perceived usefulness | “The degree to which a person believes that using a particular system would enhance his or her job performance”[25] |
| Perceived ease of use | “The degree to which an individual believes that using a particular system would be free of physical and mental effort.”[25]. |
| Social influence process | Relationships among social factors that include subjective norm, voluntariness and image[30]. |
| Subjective norm | Perceived peer pressure[30]. |
| Voluntariness | The choice to adopt the technology[30]. |
| Image | The perceived impact on one's professional image[30]. |
| Cognitive instrumental processes | Individuals evaluating how well a system or technology aligns with their job relevance, result demonstrability and output quality[30]. |
| Job relevance | The system's applicability to their work tasks[30]. |
| Result demonstrability | The visibility of benefits[30]. |
| Output quality | the performance of the system[30]. |
| Healthcare delivery | Using CARES to provide quality of care and service delivery for ART clients |
| Organizational expectations | Expectations set by the organization for using CARES |
| Continued and effective usage | Continued and effective use of CARES |
| Continuous engagement | Continuously engaging key stakeholders |
| Sustainability | the longevity and the continuous manifestation of the benefits and outcomes of digital innovations for health workers, the standard of healthcare, and patient experience long after the end of the program |

We present the results according to the TAM2 framework. As some of our findings went beyond the TAM2 framework, we introduce the additional themes of continuous engagement and sustainability. The constructs and new definitions of key terms are outlined in Table 1.

### Data Collection

HCWs took part in IDIs after 18 months of iterative CARES development and three months of NCAP field testing to optimize the CARES prototype. Data was collected at the LT clinic. The interview guide was informed by TAM. Interviews were conducted to assess their experiences with the CARES app development and implementation process, explore benefits and challenges, and identify recommendations for improvement before the broader launch of the CARES approach. All interviews were run by an external, experienced qualitative female researcher using a semi-structured interview guide (see S2 Appendix). The interviewer is an employee of the LT clinic, but was not involved with CARES. The interviewer was interested in our research and previously worked in NCAP research. IDIs were conducted in English and recorded and transcribed verbatim. The transcripts were not returned to the participants. No one else was present in the interviews besides the interviewer and respondents. The interview guide was pilot-tested with the first participant to ensure the effectiveness and clarity of the interview questions. None of the interviews were repeated, and the interviewer recorded field notes during select interviews.

### Data Analysis

We used rapid qualitative analysis to analyze the interviews. This approach streamlines the qualitative data analysis process, facilitating quicker implementation and dissemination of evidence-based innovations to reduce delays in translating research findings into practical applications [32]. Instead of employing a traditional coding tree, the interview transcripts were condensed into summary templates, utilizing data reduction techniques tailored for rapid turnaround in qualitative health services research. These summary templates were structured around domains aligned with the interview questions [33]. Two researchers initially summarized the first three transcripts and compared their summaries to standardize the template methodology [34]. The matrix was constructed by transferring the content of the summaries for each domain into an Excel file. Once all summary content was transferred, a team of researchers analyzed the matrix in a series of meetings. Insights were discussed until a consensus was reached on the themes and sub-themes. The study participants did not provide feedback on the findings.

#### Ethics

The study protocol was approved by the Malawi National Health Sciences Research Committee (#21/11/2830) and the Ethics Review Board of the University of Washington, Seattle, USA (STUDY00013936). All participants provided written informed consent.

## Results

### IDI Participants

We interviewed 15 HCWs involved with NCAP service delivery or M&E (Table 2). The majority were male (N=9) and nurses (N=10). The mean age of the participants was 38, with an average duration of employment at Lighthouse and involvement with the NCAP being seven years and four years, respectively. The interviews lasted, on average, 34 minutes.

**Table 2:** Demographic characteristics

| **Cadre** | **Female (N)** | **Male (N)** | **Median Age, years (IQR)** | **Median duration employed at Lighthouse, years (IQR)** | **Median duration involved with NCAP, years (IQR)** |
| --- | --- | --- | --- | --- | --- |
| M&E | 2 | 3 | 31(32-44) | 6(2-8) | 5(3-7) |
| Nurse | 4 | 6 | 39(32-39) | 8(3-14) | 3(2-5) |
| **Total** | **6** | **9** | **38(32-43)** | **7(2-12)** | **4(2-7)** |

IQR= interquartile range; NCAP=Nurse-led community ART

### Perceived usefulness of CARES prototype

Perceived usefulness is the extent to which a technology is expected to improve a potential user’s performance. We identified two features that contribute to the perceived usefulness of the CARES app: a) healthcare delivery and b) organizational expectations. These findings are grounded in the respondents’ expectations of the CARES app’s potential and their actual experiences in the pilot phase.

#### Usefulness of CARES prototype to improve healthcare delivery

All the IDI respondents had high expectations for CARES’ impact on improving workflow and documentation. Most respondents expected a fast and reliable device to streamline NCAP services in the community, explicitly improving data management and making things easier and faster in NCAP service delivery and M&E. Both CARES and M&E respondents highlighted that CARES was designed to be more beneficial than ODK and would reduce the nurses’ workload.

*“ the application should fasten the process of assisting the clients, we shouldn’t spend a lot of time with the client” (IDI 010)*

#### Usefulness of CARES prototype data for decision-making

In addition, nurses emphasized the CARES app’s potential to improve the quality of M&E data, including timeliness, data accuracy, and adherence calculations, mirroring EMRS functionalities. Most mentioned expectations that CARES would enhance real-time clinical decision-making when providing care. Some nurses envisaged that CARES would improve NCAP data management by reducing retrospective entry, offering valuable community data collection, and potentially freeing M&E personnel. M&E respondents echoed these sentiments, emphasizing the app’s role in bridging gaps between community-level data collection and EMRS systems.

*“ It can guide or remind us, let’s take it for instance a client needs to have viral load sample taken or the number of clients that need to be seen in a particular day. It gives us an alert on the clients that we will see…It pops up viral load for those that will be seen ” (IDI 005)*

#### Usefulness resulting from iterative HCW inputs into CARES prototype

The nurses offered extensive input on the necessary features to streamline the NCAP process based on their expectations of how CARES app should enhance their work. They shared NCAP SOPs and data collection tools and facilitated developer visits to the community to observe NCAP procedures and grasp NCAP requirements. Nearly all M&E respondents noted their participation in the development process by contributing to the data validation that enhanced the app’s flow and design so that CARES app would strengthen the data management system. Additionally, users engaged in discussions with the MoH and other external stakeholders overseeing the EMRS. During weekly virtual meetings with developers, nurses also noted that they highlighted expected features like viral load reminders and stressed the importance of syncing with the EMRS. This collaborative app development, with users informing features based on their needs, allowed users to offer insights into practical aspects and suggest needed improvements to enhance its utility. Collectively, the respondents felt that the feedback sessions were productive, believing that the app would live up to their expectations.

*We are the ones on the ground and whatever we say they* [the developers] *would understand and incorporate it. (IDI 011)*

#### Facilitators of using CARES prototype that enhanced user buy-in

All respondents stated that the CARES app helps the nurses prepare before they head to the community. Nurses praised CARES for its practicality in routine tasks, particularly liking features like viral load alerts and automatic client list generation for community visits that helped administer care, especially over the existing ODK tool. They valued CARES’ ability to potentially extract EMRS data directly, saving time spent verifying data. Moreover, some nurses noted that CARES facilitated drug management, providing the necessary quantities needed for community visits. In addition, most nurses expressed fewer data discrepancies due to EMRS data integration.

*“Previously they would do manual work to know which clients are due for viral load. Using the App, they can just generate a list of clients who are due for viral load and that makes their preparation easier and quick….. Even for drugs… this App has the drugs reconciliation per site as such the nurses will be able to know which site has run out of drugs as they are preparing where to pick the drugs, it helps in the drugs reconciliation” (IDI 002)*

M&E respondents highlighted CARES’ technical advantages over ODK, including better display of information to aid providers during a client visit, enhanced security, and reduced workload. One respondent underscored its uniqueness as there is currently no community-based ART management tool.

*“It [my workload] has been reduced …previously the process of printing will take close to two hours, that is just to pull the data and print. This time when they are back, we just pull the data and print within a few minutes” (IDI 007)*

Furthermore, the M&E respondents praised CARES’ design for its compatibility with the EMRS’s data schema, ensuring consistent data collection for M&E purposes and data security due to encryption.

#### Barriers using the CARES prototype that discouraged continued uptake

All IDI respondents consistently voiced dissatisfaction regarding the speed of the CARES app performance, which worsened the nurses’ workload and increased the time spent with the client waiting for the app to load. Respondent IDI 15 complained that “*one of the major frustrations for the nurses, the expectation was that they will be able to see a lot of people within a short period using the app, but it is not helping in that area* “. They also highlighted that the app has not yet achieved integration with the EMRS—a key anticipated feature. As a result, they still have to manually input data into the EMRS.

All nurse respondents detailed concerns about the slow performance leading to manual data entry, thereby undermining its intended efficiency. Most nurse respondents noted discrepancies in CARES’ data, including duplicates and client information errors. In addition to the data discrepancies and speed, the M&E respondents were concerned with compromising the NCAP database integrity if wrong information was entered into the app. They also reported missing client records, who had to be re-enrolled in the application. In addition, they stated that they need features like daily client schedules and filter options for support groups to ease their workload. As a result, the respondents agreed that CARES is not superior to ODK in its current state, lamenting that the “*CARES app aimed at shortening the time taken to see the client and data entry… but the process is still the same”(IDI 008)*.

#### Organizational Expectations for CARES Prototype Testing

We identified organizational expectations as an influence on the perceived usefulness of the CARES app. Healthcare providers’ use of the app, even when it was not performing to expectations, appears driven by the expectations set by the research teams and the CARES developers. For instance, one nurse reported that due to the app becoming too slow, some nurses chose not to use CARES in the community while providing care as initially intended. Instead, to meet organization expectations and requirements for CARES testing, they opted to enter data into the app upon returning to the facility. IDI noted that they did not use CARES in the community, but still felt obligated to, *“enter the data not for our* [NCAP] *sake but for them [developers] to see that we have entered the data” (IDI 008)* . Forging CARES when it lagged while in the field and reverting to ODK for later entry into CARES created a burdensome workload for CARES providers and M&E teams that met requests for research needs on top of care delivery.

### Perceived ease of use for CARES prototype

Perceived ease of use is defined as the degree to which a person believes that using a particular system would be free from effort. Several factors affected CARES’ perceived ease of use. These included having an interface design similar to the EMRS, making it more user-friendly, and using encryption for safer synchronization with the EMRS.

However, perceived ease of use was overshadowed by the slowness of the app and its tendency to reset. All highlighted poor usability issues related to the app’s speed and navigation.

*“We are unable to use the app because it is slow. When I say we end up releasing the client that means we use the old approach where we would capture the information manually and enter EMRS when we are back, as it was before CARES” (IDI 003)*

M&E respondents noted the app’s workflow challenges, such as the need for a streamlined data entry process. Additionally, the lack of an EMRS sync feature limited the extent of its utilization, increasing rather than decreasing the current workload. Similarly, the nurses expressed frustration with the app’s slow performance, citing instances where delays hindered their ability to collect and enter client data efficiently during field visits. Navigating from one CARES task or care page to another care component was a repeated source of frustration.

*“CARES has sections/pages, let’s say reception and after that you send the information, so instead of the system to continue from reception to vitals, it requires one to search for the client again once that page is sent… so you must search for the client again. You may search for the client maybe 7 times for you to complete the tasks from reception to next appointment” (IDI 013)*

### Potential for continued and effective use of CARES

In our study, given that all respondents have had experience with the CARES prototype, we framed the ’Intention to Use’ within the context of ’Intention to Continue Using CARES’.

### Optimism that CARES can be improved

Despite the extensive challenges with the CARES prototype, the IDI respondents universally expressed optimism and faith in its potential. Some respondents emphasized the importance of attitude for CARES’ success, highlighting the need for a mindset shift towards accepting the app. They stated that with this change, CARES would thrive. Additionally, the active participation of respondents in the development process indicates their involvement and investment in the app’s success, which can also impact their willingness to continue using it, as noted by IDI 10, who stated, “*We have been so patient because we know where we are coming from.”* Some respondents hesitated to express negative opinions about the app because it is still in the beta version, and they remained hopeful that further improvements could be made. Other factors that may influence the intention to continue using CARES include mandatory compliance encouraged by organizational leadership.

### Themes beyond TAM

While the TAM provided a valuable framework for understanding the respondents’ acceptance of the CARES prototype based on perceived usefulness and perceived ease of use, not all themes emerging from the interviews could be neatly categorized within TAM constructs. We identified additional themes that influence the continued use of the CARES prototype, delving into the long-term maintenance and sustainability of CARES.

### Continuous engagement

We observed that continuous engagement is vital to the continued use of CARES. This involves capacity building and engaging key external stakeholders to address challenges and maintain the system.

#### Capacity building

The M&E respondents highlighted the need for comprehensive training and skill transfer for sustainable app maintenance. Nurses added that extended observation for enhancements and broader training, including other cadres such as pharmacists, for effective NCAP support.

[Training for CARES should include] *what is the CARES, what do we want to achieve in terms of the objectives, and what improvements are we intending to achieve by the end of the day. CARES, the providers should understand the objective which is retaining people in care (IDI 002)*

#### Stakeholder engagement

Stakeholder engagement was emphasized primarily by M&E respondents, who stressed the need for ongoing collaboration between CARES developers and MoH’s HIV/AIDS and Digital Health Unit to align the app with national standards and address mHealth gaps. One respondent emphasized the importance of technical expertise:

“*I think we also need some kind of technical expertise on the use of the system, especially when you need pulling and pushing data between EMR and the CARES application” (IDI 009).* In addition, they deemed concurrence from the MoH, and other key stakeholders involved in the Malawi Health Information System as vital, particularly for integrating CARES with the EMRS.

### Sustainability

Sustainability emerged as a theme that influenced the continued use of CARES. This included a focus on capacity building, stakeholder engagement, infrastructure, data security, and integration with other services.

#### Infrastructure

IDI respondents underscored the need for resources to maintain CARES, such as tablets for nurses, drug storage, and adequate human resources, as critical for sustainability. Furthermore, the IT manager, stressed the need for proper infrastructure, like servers and support staff, for scaling up.

The data manager suggested appointing an organization like Lighthouse as the app’s custodian if CARES was scaled to other facilities beyond Lighthouse.

*If there could be a deliberate plan that the one who was given the custodian of the application should continue with the system, then it will be good…there is no capacity of government to use or maintain the system. (IDI 009)*

#### Data Security

Respondents unanimously identified data security as paramount to strengthening CARES, advocating for password protection, unique user IDs, and internet access controls in the CARES app. Further suggestions included short data retention within the app, with local clinic storage post-EMRS sync. They also recommended using biometric login, data wipe protocols, clear tablet security responsibilities, and individualized app access to ensure data integrity and prevent misuse.

*It should also have a feature that the moment you make mistakes to access the CARES app in case you have made the mistake on the log in details …then the device should wipe all the data. (IDI 008)*

#### Integration with other services

When asked about suggestions for expanding CARES prototype features, they proposed enhancements such as monitoring viral load orders, transitioning to online platforms, and adding dashboards to expand CARES functionality. Some nurses recommended expanding the app’s services to incorporate other healthcare services, such as cervical cancer screening and non-communicable diseases.

*If clients are already available in the app, then they can bring a scanner instead of searching for the client, we can scan the barcode and it will be able to bring the client, that will help to reduce the amount of time. (IDI 006)*

The integration of CARES with other mHealth interventions, notably the Two-way Texting (2wT) SMS delivery system currently piloted at Lighthouse, was also recommended by some M&E respondents.

*Maybe if it could be integrated with their phones, maybe getting the reminder direct from the CARES to the client, the appointment, getting medication, motivation…Just as the way it is with two-way texting. (IDI 009)*

## Discussion

This study analyzed healthcare providers’ perspectives on the mHealth CARES prototype, examining its role in facilitating NCAP services for clients, nurses, and M&E teams. We identified important factors facilitating HCWs’ acceptance of mHealth technologies— healthcare delivery improvement, organizational expectations, collaborative development, and sustainability initiatives - and formidable barriers to CARES success, including app performance issues, data discrepancies, navigation challenges, data security concerns, infrastructure, and resource limitations. We framed our results based on the extended TAM[30] that prioritizes perceived usefulness and perceived ease of use. To these, we added additional themes-stakeholder engagement and sustainability. We discuss how these results informed additional CARES iterations, improvements, and plans for future adaptations.

First, applying the TAM2 framework helped underscore the influence of perceived usefulness and ease of use in CARES technology adoption. HCWs anticipated the CARES prototype would enhance efficiency, aligning with TAM2’s job relevance construct [30]. They praised CARES‘ ability to streamline tasks such as community visit preparations and drug management, indicating high perceived usefulness, but performance challenges negatively impacted the perceived ease of use. Additionally, the concept of subjective norm within TAM2 [30] reflects the societal influences on an individual’s decision to use a technology. Providers continued using CARES amidst greater workloads, showing their commitment during its prototype phase and underscoring the role of stakeholder expectations in technology adoption decisions. It suggests that when HCWs believe in the importance of the specific technology to their supervisor’s or the organization’s goals, they may be more willing to adapt their work practices to incorporate the technology at the development stage.

Moreover, we identified user optimism, continuous engagement, and sustainability as crucial for the CARES app’s continuing and effective use, confirming past research that links HCW attitudes and stakeholder commitment to mHealth success [35–37]. On the positive side, providers accepted many of the current CARES weaknesses and remained optimistic about continuing to use CARES if improvements were attained. Nurse users emphasized the potential for a positive impact on patient care, while M&E respondents focused on technical and programmatic considerations. These improvements could increase healthcare quality as a result of the mHealth tool, a facilitator of mHealth implementation success, as seen previously [35–38]. However, similar to the findings from a systematic review of HCW acceptability of mHealth interventions, we found that continued issues in CARES technical failures and resulting increased workload were significant barriers to CARES, curbing enthusiasm[39]. HCWs reported frustration about the CARES prototype’s slow performance and difficulties with EMRS synchronization, again decreasing engagement and buy-in. HCW users cited the unfilled promise of improved workflow, streamlined data management, and overall efficiency for NCAP service delivery but also suggested that overcoming these issues would further positive attitudes toward future CARES interactions. Lastly, sustainability, which centered around critical improvements in infrastructure, connectivity, data security, and technical support, emerged as essential to scaling up and sustaining mHealth intervention. Sustainability appeared tied to a strong commitment by both HCWs and relevant stakeholders, as seen elsewhere in mHealth expansion success regionally [40,41].

The HCD process appeared crucial for HCWs’ buy-in. Their involvement in the design phase, sustained through an 18-month engagement alongside their duties, reflects an enormous dedication to NCAP clients and the improvement of care delivery. Moreover, their consistent inputs for iterative testing helped increase CARES‘ prototype tailoring to the Malawian context and the unique needs of NCAP and the EMRS at Lighthouse. This approach also secured organizational backing, addressing a common barrier in mHealth implementation [37,42] . These issues necessitated supplementary documentation methods, raising questions about the app’s practicality and, thus, the usefulness of the app, even if they recognize its theoretical benefits [43,44]. Technological challenges, which are common barriers to implementing and adopting mHealth interventions [40,45,46] were among the factors contributing to the delays in addressing CARES issues.

Our research underscores that while the TAM provides a robust framework for understanding the key determinants of technology acceptance, it must be contextualized within healthcare professionals’ specific operational workflows and user experiences. Although TAM and TAM2 were helpful frameworks, we could not apply all aspects of the theory due to the timing of the IDIs within the CARES prototype testing timeframe. We excluded behavioral intention from our model because, in our study, participants were interviewed after already experiencing the intervention, describing perceptions of actual usage rather than intentions. Previous research on technology acceptance showed that behavioral intention becomes a less significant predictor of actual use when the likelihood of use is initially high [47]. However, we also included the continued use of CARES in our model to explain if and how HCWs will continue using CARES in the future.

Our findings highlight the importance of user-centered design in healthcare app adoption and efficacy where HCW and stakeholders are involved in every step—from design to iterative testing—to tailor the app to their actual needs and preferences. Additionally, securing organizational support and aligning the app with institutional objectives encourages healthcare workers to seamlessly incorporate new technology into their daily workflow.

The study faced several limitations: Firstly, the sample size was small and consisted of deliberately selected individuals. However, all providers involved in NCAP were interviewed thus incorporating all individuals relevant to the research. Secondly, the evaluation occurred while the app was still undergoing testing and enhancements, suggesting its usability might evolve over time. Thirdly, the interviewer’s affiliation with Lighthouse Trust and familiarity with NCAP may have influenced participant responses, although she had no direct involvement with CARES. Lastly, despite efforts to encourage candid feedback, social desirability bias may have influenced participants’ responses, potentially skewing the findings related to the CARES app.

## Conclusion

Overall, HCWs largely embraced the promise of the CARES prototype even while recognizing that the reality of current CARES falls short. High levels of HCW involvement in the app’s development likely facilitated their commitment to CARES success. The insights obtained from these interviews offered guidance for refining the CARES app, and these recommendations are being systematically addressed through iterative CARES improvements. If incorporated successfully, continued user optimism for CARES‘ impact to streamline their work and improve patient care will be rewarded. In contrast, if the CARES prototype is not enhanced for a smoother user experience in the community and clinic, and integration with the EMRS is not achieved to streamline the data delivery workload, there is a high risk of discontinuation. Continued collaboration among developers, NCAP providers, and other stakeholders, paired with sufficient financial investment in CARES improvement, is the essential next step. With that support, CARES can realize its potential to extend the reach of the EMRS into community settings, ensuring that clients in clinic and community settings receive high-quality, integrated care.

## Supporting Information

**S1 Appendix :** COREQ (COnsolidated criteria for REporting Qualitative research) checklist

**S2 Appendix :** Interview Guide

## Data Availability

Interested researchers may reach out to the Human Subjects Division contact for our study, to request access to the datasets. Access to de-identified interview transcripts, de-identified demographic data, and codebook will be restricted to those who complete data sharing agreements.

## Acknowledgments

The research reported in this publication was supported by the National Institutes of Health (NIH), the National Institute of Mental Health (NIMH) under award number R21MH127992 (“The Community-based ART REtention and Suppression (CARES) App: an innovation to improve patient monitoring and evaluation data in community-based antiretroviral therapy programs in Lilongwe, Malawi”), and multiple principal investigators (CF and HT).

The authors would like to thank all the healthcare workers who shared their opinions and experiences in the interviews, the Lighthouse Trust community health services program, and the research department for their partnership in implementing the CARES application.

## Author Contributions

**Conceptualization:** CF, HT

**Data Curation:** OS

**Formal Analysis:** CKK,ABR

**Funding Acquisition:** CF, HT

**Investigation:** OS, CKK, ABR, GO, CF

**Methodology:** CKK, ABR, CF

**Project Administration:** JH, AT

**Supervision:** CF

**Validation:** CKK, ABR, GO, CF

**Visualization:** CKK

**Writing:** Original Draft Preparation_CKK

**Writing Review & Editing**: CKK, ABR, GO, WG, MP, CF

